# Characterization of highly pathogenic avian influenza virus in retail dairy products in the US

**DOI:** 10.1101/2024.05.21.24307706

**Authors:** Erica Spackman, Deana R. Jones, Amber M. McCoig, Tristan J. Colonius, Iryna Goraichuk, David L. Suarez

## Abstract

In March 2024 Clade 2.3.4.4b H5N1 highly pathogenic avian influenza virus (HPAIV) was detected in dairy cattle in the US and it was discovered that the virus could be detected in raw milk. Although affected cow’s milk is diverted from human consumption and current pasteurization requirements are expected to reduce or eliminate HPAIV from the milk supply, a study was conducted to characterize whether the virus could be detected by quantitative real-time RT-PCR (qrRT-PCR) in pasteurized retail dairy products and if detected, to determine whether the virus was viable. From April 18 to 22, 2024 a total of 297 samples of Grade A pasteurized retail milk products (23 product types) were collected from 17 US states and represented products from 132 processors in 38 states. Viral RNA was detected in 60 samples (20.2%) with titer equivalents of up to 5.4log_10_ 50% egg infectious doses (EID_50_) per ml, with a mean and median of 3.0log_10_/ml and 2.9log_10_ /ml respectively. Samples that were positive for type A influenza by qrRT-PCR were confirmed to be clade 2.3.4.4 H5 HPAIV by qrRT-PCR. No infectious virus was detected in any of the qrRT-PCR positive samples in embryonating chicken eggs. Further studies are needed to monitor the milk supply but these results provide evidence that infectious virus did not enter the US pasteurized milk supply before control measures for HPAIV were implemented in dairy cattle.

**Importance:** Highly pathogenic avian influenza virus (HPAIV) infections in US dairy cattle were first confirmed in March 2024. Because the virus could be detected in raw milk a study was conducted to determine whether it had entered the retail food supply. Pasteurized dairy products were collected from 17 states in April 2024. Viral RNA was detected in 1 in 5 samples but infectious virus was not detected. This provides a snap-shot of HPAIV in milk products early in the event and reinforces that with numerous safety measures, infectious virus in milk is unlikely to enter the food supply.

## Introduction

Cow’s milk and milk products are an important source of nutrition for humans. In the US, “Grade A” milk is regulated by a federal-state partnership, the National Conference on Interstate Milk Shipments (NCIMS), and is administered through adopted regulations, the Pasteurized Milk Ordinance (PMO) (https://www.fda.gov/media/140394/download). The NCIMS helps the industry produce a safe and wholesome product for the consumer. This regulatory system has multiple layers to ensure food safety. Cows with mastitis and other disease conditions that could affect milk quality and safety are milked separately, and the abnormal milk is not included in the supply for human consumption. Milk is also typically picked up from the farm at regular intervals, and the bulk milk (milk pooled from 600-700 cows) is routinely tested for commonly used antibiotics and other substances before pasteurization (https://www.fda.gov/food/food-compliance-programs/national-drug-residue-milk-monitoring-program). Samples are also analyzed on a recurring basis for somatic cell and bacterial plate counts to monitor quality management practices.

Pasteurization is another pivotal layer of the federal-state milk safety system. The primary method for pasteurization of fluid milk is typically through a continuous flow pasteurizer by high temperature short time; 72°C for 15 seconds is the most used approved method by regulation in the US according to the PMO. Variations in pasteurization time and temperatures are allowed that achieve the same goal of killing pathogenic bacteria and to reduce spoilage bacteria that will in effect increase the shelf life of the milk. The milk is then packaged and sent to retail markets with strict temperature controls that further ensures the safety and quality of the product.

Infection of dairy cattle with clade 2.3.4.4b H5N1 highly pathogenic avian influenza virus (HPAIV) was first reported in the US on March 25, 2024 (1). Diagnostic testing of milk from the initial cases detected viral RNA by real-time RT-PCR. The potential for HPAIV to enter the food supply is believed to be mitigated because symptomatic cows have decreased milk quality and production thus preventing the milk from entering the food supply due to milk safety controls. Poor quality milk is normally diverted from the milk supply for human consumption. However, because HPAIV has never been described in dairy cattle, milk has not been monitored for the virus.

Historically, documentation of influenza A virus infection in cattle has been sparse with only a few reports of clinical disease (2-4), and there has not been evidence of sustained transmission among cows (5). More recently, serologic studies on respiratory disease or drops in milk production were reported in Northern Ireland that were associated with a rise in convalescent antibody titers to influenza A subtypes that are consistent with human seasonal influenza but no virus was isolated to confirm the lineage present (3). Several experimental studies from the 1950s clearly show that the direct inoculation of the human PR8 influenza A virus strain or Newcastle disease virus into the udder of lactating dairy cows or goats could result in infection with measurable virus shedding, however, the studies did not describe clinical disease or mastitis in the challenged animals (6-9). Until the recent outbreak of clade 2.3.4.4b HPAIV in dairy cattle with sustained transmission, infection of bovines with type A influenza was not previously reported and therefore was not considered to be an important pathogen of cattle which delayed initial recognition of the infection.

Because the clade 2.3.4.4b H5 HPAIVs belong to the goose/Guangdong/1996 H5 HPAIV lineage, which is known to have zoonotic potential (10), the objective of this study was to screen pasteurized retail dairy products for the presence of viral RNA. Positive samples were subsequently evaluated for the presence of live virus in embryonating chickens eggs. Importantly, human infections with clade 2.3.4.4 H5 HPAIV are rare and numerous risk assessments have concluded that the risk to the general public is very low (https://www.ecdc.europa.eu/en/infectious-disease-topics/z-disease-list/avian-influenza/threats-and-outbreaks/risk-assessment-h5, https://www.who.int/publications/m/item/assessment-of-risk-associated-with-recent-influenza-a%28h5n1%29-clade-2.3.4.4b-viruses, https://www.fao.org/animal-health/situation-updates/global-aiv-with-zoonotic-potential/en).

## Results

### Virus detection

A total of 297 samples representing 23 pasteurized dairy product types (Supplementary Table) were collected from 17 states which represent products produced at 132 processing locations in 38 states. Of these, 20.2% (60/297) were positive for the detection of influenza A RNA by qrRT-PCR (Table 1). Virus titer equivalents for positive samples ranged up to 5.4log_10_ 50% egg infectious doses (EID_50_) per ml, with a mean and median of 3.0log_10_/ml and 2.9log_10_ /ml respectively (Supplementary Table). Fluid milk with different fat contents represented 64.0% (n=190) of the products tested and 75% (n=60) of the samples in which influenza A was detected by qrRT-PCR.

**Table 1.** Detection of influenza A in pasteurized retail dairy products by quantitative real-time RT-PCR. Titer values are expressed as log_10_ 50% egg infectious doses determined by a standard curve using quantified virus. No infectious virus was detected in any of the qrRT-PCR positive samples.

| **Product** | **# positive / total tested**<br>**(% positive)** | **Mean titer equivalents**<br>**(± standard deviation)** |
| --- | --- | --- |
| Whole milk | 16/68 (23.5) | $3.0 \pm 1.1$ |
| 2% reduced fat milk | 16/58 (27.6) | $3.1 \pm 1.2$ |
| 1% low fat milk | 9/28 (32.1) | $3.1 \pm 1.2$ |
| Skim milk | 4/36 (11.1) | $3.3 \pm 0.7$ |
| Half and half | 6/25 (24.0) | $2.3 \pm 1.0$ |
| Yogurt | 0/14 (0) | Not applicable |
| Cream | 3/17 (17.6) | $2.3 \pm 0.9$ |
| Cottage cheese | 1/21 (4.8) | $2.6 \pm 0.0$ |
| Sour cream | 5/30 (16.7) | $3.4 \pm 1.2$ |
| Total | 60/297 (20.2) | $3.1 \pm 1.1$ |

A subset of the samples positive for type A influenza by qrRT-PCR (n=30) were confirmed to be clade 2.3.4.4 HPAIV by a lineage specific qrRT-PCR test; 100% (30/30) were positive.

A total of 60 samples that were positive for type A influenza were tested for infectious virus by standard testing in ECE. Infectious virus was not detected in any samples (Supplementary Table).

## Discussion

In March 2024 HPAIV was discovered in the milk of infected dairy cattle in the US. Samples were collected from retail markets in April 2024 to assess a variety of products to provide data for an initial safety risk assessment of the national milk supply. Samples were selected to be representative of dairy processors in states that have confirmed HPAIV infected dairy cattle, and states that have not reported infected herds. Of note, due to the complexity of the milk distribution system, the location of where milk was processed may not correlate with the location where the milk was produced. Commercial milk is typically pooled from several dairy farms and routed for bulk processing (i.e., pasteurization) and distribution to multiple states is a common industry practice. For example, a product could have been produced by cows in one state, then processed in a different state, and then sold commercially in a third state.□

Most importantly, although viral RNA was detected by qrRT-PCR in 20.2% of the samples, no infectious virus was detected by testing for replication in ECE, which is a highly sensitive bioassay for avian influenza virus detection (11, 12). Positive qrRT-PCR indicates that some viral RNA entered the milk supply, however, it can’t be determined at what stage, if any, the virus was infectious. First, cows rapidly develop antibodies after infection which are present in milk and will inactivate the virus. Second, virus is inactivated by pasteurization and possibly the high shear force of homogenization. Work with continuous flow pasteurization is in progress to confirm the conditions for virus inactivation.

This study has several limitations that make wider extrapolation of HPAIV RNA levels in pasteurized dairy products difficult. First, the sample size is small. The scope of this study was to obtain an initial snap-shot of whether dairy products had evidence of virus in retail milk samples after the detection of virus in raw milk from dairy cows. Further, some samples were intentionally collected from regions with known HPAIV infected dairy herds, therefore these data likely provide a higher positivity rate than would be expected from a random testing process. Since the recognition of dairy cattle infection with HPAIV, farmers are more aware of the disease, and diagnostic testing can occur in many of the USDA approved laboratories in the National Animal Health Laboratory network. Currently, dairy cattle must be tested before moving across state lines (https://www.aphis.usda.gov/sites/default/files/dairy-federal-order.pdf) helps mitigate contaminated milk from entering the human food supply. Finally, regardless of the specific detection of HPAIV infection, cows will develop mastitis which will also result in removing their milk from the food supply.

In general, numerous measures in the milk production process will greatly reduce, if not eliminate, the risk for infectious influenza A virus entering the retail milk supply. First, approximately 99% of the US commercial milk supply (https://downloads.usda.library.cornell.edu/usda-esmis/files/4b29b5974/hq37xb74r/s1786b07q/mlkpdi24.pdf) that is produced on dairy farms in the US comes from farms that participate in the Grade “A” milk program and follow the PMO (https://www.fda.gov/media/140394/download), which includes numerous layers of quality controls that help ensure the safety of dairy products. Second, the US federal-state milk safety system requires that milk from sick cows is diverted for further processing or is destroyed.

More studies are needed to characterize the risk of HPAIV entering the milk supply long term but this study provides initial evidence that infectious HPAIV has not reached the US retail milk supply. A combination of the previously implemented sanitary control measures (e.g., PMO) and new HPAIV specific measures are expected to further ensure a safe milk supply.

## Materials and Methods

### Retail dairy product sample collection

The US Food and Drug Administration (FDA) collected 297 samples at retail locations in 17 states between April 18 and 22, 2024. Sample sites were selected by local FDA Milk Specialists and field staff, in the Office of Regulatory Affairs. Samples were shipped directly by overnight courier to the US National Poultry Research Center, USDA-Agricultural Research Service where testing was conducted. Sample collection was designed to include both products processed in states where HPAIV infections in dairy herds had been confirmed by the National Veterinary Services Laboratories, APHIS-USDA, at the time of collection, as well as samples from states with no confirmed infections in dairy herds. Within these bounds, sample collection was random and based on retail availability. Samples represented pasteurized retail dairy products produced at 132 processors in 38 states (AR, AZ, CA, CO, CT, FL, GA, IA, ID, IL, IN, KS, KY, MA, ME, MI, MN, MO, NC, ND, NE, NH, NJ, NV, NY, OH, OK, OR, PA, SC, TN, TX, UT, VA, VT, WA, WI, WV). Samples included fluid milk (whole, 1%, 2%, skim), cream (heavy cream, light cream, and similar), half & half, cottage cheese (and similar), sour cream, and yogurt (Supplementary Table). All samples were Grade A pasteurized dairy products regulated under the PMO. (https://www.fda.gov/media/140394/download, https://www.fda.gov/food/guidance-documents-regulatory-information-topic-food-and-dietary-supplements/milk-guidance-documents-regulatory-information) by FDA and its state milk regulatory partners.

#### Sample processing

Samples were immediately processed after receipt. Product with temperatures >7°C were discarded and are not included in the sample numbers of this study. Samples were assigned a unique accession number and the original packaging was labeled and stored at 4°C. Product origin (US state) and product type were recorded.

Approximately 50ml of each product was portioned into sterile containers. Each sample was processed for RNA extraction and quantitative real-time RT-PCR (qrRT-PCR) as described below. Positive samples with titer equivalents of ≥ 3.9log_10_ 50% egg infectious doses (EID_50_)/ml based on qrRT-PCR were quantified in embryonating chicken eggs (ECE) and samples with titers ≤3.8log_10_ EID_50_/ml were tested for viable virus in ECE as described below. The cut-off for quantification was selected because it was expected that, if present, the quantity of infectious virus would be lower than the quantity detected by qrRT-PCR and quantification of low levels would not be informative.

#### RNA extraction

RNA was extracted from fluid homogenized dairy products using the MagMax magnetic bead extraction kit (Thermo Fisher Scientific, Waltham, MA) in accordance with manufacturer’s instruction. Semi-solid products (e.g., sour cream, yogurt, cottage cheese) were extracted using a hybrid procedure with Trizol LS (Thermo Fisher Scientific) and the MagMax magnetic bead kit. Semi-solid products were portioned by spatula based on weight (approximately 0.25g). Briefly, VetMAX Xeno (Thermo Fisher Scientific) was used as an extraction and internal positive control was added to the Trizol LS for each reaction prior to sample addition. Then 0.25ml or 0.25g of product was added to 0.75ml of Trizol LS and mixed. The mixture was incubated at room temperature for 7-10minutes and 0.2ml of chloroform was added and mixed, incubated at room temperature for an additional 7-10minutes and centrifuged for 10minutes at 15,000xg at 4°C. RNA was recovered from 0.05ml of the aqueous phase by the MagMax magnetic bead kit in accordance with the kit instructions.

#### Quantitative real-time RT-PCR

A qrRT-PCR test targeting the influenza A M gene was run on a QuantStudio5 (Thermo Fisher Scientific) as described (13). The primers and probe for the internal control were used as directed by the kit instructions. Titer equivalents were determined by including a standard curve derived from RNA extracted from a 10-fold dilutions series of quantified avian influenza virus stocks (14). A subset of the influenza A qrRT-PCR positive samples were tested with and additional qrRT-PCR test that is specific for the 2.3.4.4b H5 lineage with a highly pathogenic cleavage site (15).

#### Virus detection and quantification in embryonating chickens eggs

All samples (1ml) were treated for 1hr at ambient temperature (approximately 21°C) with antibiotics (final concentration: penicillin G 1000 IU/ml, streptomycin 200 μg/ml, gentamicin100 μg/ml, kanamycin 65 μg/ml, amphotericin B 2 μg/ml). Then dilutions were made in brain heart infusion (BHI) broth with antibiotics for samples that were quantified. Semisolid samples were mixed 1:1 (0.5g:0.5ml)) with brain heart infusion broth prior to inoculation into ECE or dilution. Samples were inoculated for virus detection (undiluted for 2 passages) or quantified using standard methods (16, 17). Hemagglutination assay was used to confirm the presence of avian influenza virus (18).

## Data Availability

All data produced in the present work are contained in the manuscript and supplemental data.

## Acknowledgements

The authors gratefully thank Nathan Anderson, Frankie Beacorn, Tarrah Bigler, Nick Chaplinski, Suzanne DeBlois, Edna Espinoza, Jesse Gallagher, Javier Garcia, Jessica Gladney, David Haley, Anne Hurley-Bacon, Lindsay Killmaster, Scott Lee, Stephen Norris, David Pearce, Timothy Roddy, Melinda Vonkungthong, Stephen Walker, Robin Woodruff, and Ricky Zoller for technical assistance with this work.

Any use of trade, firm, or product names is for descriptive purposes only and does not imply endorsement by the US Government. This research was supported by US Department of Agriculture (USDA)-Agricultural Research Service Project No. 6040-32000-081-00D and the US Food and Drug Administration by IAA contract number pending. All opinions expressed in this paper are the authors’ and do not necessarily reflect the policies and views of the USDA or FDA. The USDA is an equal opportunity provider and employer.

## Notes

### Competing Interest Statement

The authors have declared no competing interest.

### Funding Statement

This study was funded by the US Dept of Agriculture project # 6040-32000-081-00D and the US Food and Drug Administration project # pending.

